# Geranylgeraniol supplementation leads to an improvement in inflammatory parameters and reversal of the disease specific protein signature in patients with hyper-IgD syndrome

**DOI:** 10.1101/2024.07.17.24309492

**Authors:** Anna Sediva, Martin Orlicky, Petra Vrabcova, Adam Klocperk, Tomas Kalina, Hideji Fujiwara, Fong-Fu Hsu, Monika Bambouskova

## Abstract

Mevalonate kinase (MVK) deficiency, a rare autosomal recessive disease, significantly impacts metabolism and immunity, leading to mevalonic aciduria in severe cases and hyper-IgD syndrome (HIDS) in partial deficiency. These conditions arise due to disruptions in the mevalonate pathway, which is essential metabolic pathway responsible for the synthesis of non-sterol isoprenoids and other molecules. The resulting metabolic blockade triggers autoinflammatory responses, primarily due to deficient isoprenoid intermediates such as geranylgeranyl pyrophosphate (GGPP). This first reported pilot study evaluates the safety and efficacy of dietary geranylgeraniol supplementation (GG) in three patients with HIDS. Over three months, GG supplementation showed no liver toxicity and did not alter lipid profiles. Although GG did not rise the plasma levels of GGPP, the plasma proteomics showed significant changes induced by GG. Proteomic analysis further revealed that GG supplementation can reverse some of the features of HIDS-specific plasma protein signature, highlighting its potential to modulate inflammation and protein prenylation pathways. These findings suggest that GG supplementation could be a promising metabolic intervention to mitigate inflammation in HIDS, warranting further, more targeted investigation in larger clinical trials.

## Introduction

Mevalonate kinase (MVK) deficiency is a rare autosomal recessive disease that falls into the area of inborn errors of metabolism, but also into inborn errors of immunity due to the consequences the metabolic defect exerts on the immune system. A profound MVK deficiency manifests with mevalonic aciduria, a severe condition characterized by inflammatory episodes, syndromic and neurological impairments, and psychomotor retardation. In contrast, milder deficiency with partially preserved MVK function leads to hyper-IgD syndrome (HIDS). The metabolic disturbance is a direct consequence of blockade in the mevalonate pathway, which is crucial for production of isoprenoids and sterols that are key for synthesis of various biologically significant molecules including cholesterol, dolichol, and ubiquinone. The immunological consequences of MVK deficiency manifest as an autoinflammatory disease accompanied by reccurent febrile episodes and elevated serum IgD levels, a common, albeit inconsistent marker from which HIDS gets its name^1,2^.

The autoinflammatory state associated with MVK deficiency is linked to isoprenoid deficiency due to the blockage in the metabolic pathway. Isoprenoids are essential for protein prenylation, a post-translational modification attaching 15-carbon (farnesyl) or a 20-carbon (geranylgeranyl) isoprenoid lipid to the Cys residue by specific transferases. Large group of proteins regulated by prenylation are small GTPases such as Ras, Rab, Rho and others^3^. Attached prenyl groups can regulate cellular localization and function of the proteins and prenylation was found to be defective in patients with HIDS^4^.

The inflammatory manifestations in HIDS are primarily mediated by inflammasome activation, which likely occurs through several distinct mechanisms. The deficiency of isoprenoids, particularly geranylgeranyl pyrophosphate (GGPP), leads to inactivation of RhoA GTPase, which subsequently triggers the pyrin inflammasome^2,5,6,7^. Additionally, defective protein prenylation, particularly in small GTPases, also activates the NLRP3 inflammasome in monocytes^8^. These findings suggest that both pyrin and NLRP3 inflammasomes participate in inflammation induced by MVK dysfunction, with isoprenoid deficiency serving as a crucial trigger of these inflammatory processes.

The accumulation of mevalonate upstream of the metabolic block in the mevalonate pathway may also contribute to the condition. The mevalonate buildup in monocytes from patients with HIDS resulted in their inflammatory state and a phenomenon known as “trained immunity”. This associates with an enhanced non-specific inflammatory immune response, which can further contribute to inflammation in the context of MVK deficiency^9^.

Apart from its complex effects on innate immunity, MVK deficiency, primarily through reduction of isoprenoids such as GGPP, also influences adaptive immunity. A recent study highlights significant role of GGPP in B lymphocytes connected to the defective IL-10 secretion in patients with MVK deficiency. Therefore, reduced B cell-derived anti-inflammatory IL-10 production may be a contributing factor to the inflammatory phenotype in HIDS^10^. Furthermore, mevalonate metabolism and protein prenylation were also shown to be involved in the regulation of T cell homeostasis; in this context protein geranylgeranylation exhibits unique role in enabling thymic egress, while farnesylation is more involved in the regulation of peripreral T cell homeostasis^11^.

Another intriguing aspect of the MVK deficiency is the elevated level of IgD, which gives rise to the name “hyper-IgD syndrome.” Although the increase of IgD is also observed in other autoinflammatory diseases, it is most pronounced in HIDS. The mechanism behind the elevated IgD levels remains unclear. Recent studies suggest that the increase in IgD may be an independent phenomenon, possibly not directly responsible for the inflammation^12^. IgD, along with IgM, serves as a membrane receptor involved in B lymphocyte development and activation. IgD and IgM are expressed on B lymphocytes in specifically arranged membrane islands with a distinct nanostructure^13^. A recently published hypothesis provides a new perspective on the potential cause of elevated IgD, linking it to a reduction in signaling through membrane IgM. Unlike signaling through IgD, signaling through IgM requires MAPK activation which depends on protein prenylation^14,15^. According to this hypothesis, the increased IgD level in MVK defficiency is a result of disrupted balance in IgM/IgD expression and signaling on B lymphocytes due to impaired MAPK pathway. This signaling imbalance is caused by impaired prenylation of components in the MAPK signaling cascade, particularly Ras^14^.

While this hypothesis remains to be fully validated, shortage of isoprenoids leading to impaired protein prenylation due to the mevalonate pathway blockade has been repeatedly demonstrated and recognized as a crucial factor in the inflammatory manifestations of the MVK deficiency. The rationale for supplementing isoprenoids to restore protein prenylation is supported by numerous *in vitro* and *in vivo* studies. Specifically, GGPP or its derivative geranylgeraniol, was shown to restore prenylation-associated defects in cells with disfunctional MVK^8,10,16^ and reduced inflammation in mouse model of chemically induced MVK deffcicency^17^. This strategy is consistently highlighted in publications as a plausible approach to rectify the metabolic defect caused by MVK dysfunction, however its potential to treat HIDS in human has not yet been tested.

In light of the aforementioned evidence, we conducted a pilot study evaluating the effects of dietary supplementation of geranylgeraniol in a small group of patients with MVK deficiency presenting with HIDS.

## Patients and methods

### The pilot study

In the pilot study, we aimed to verify the safety and efficacy of the dietary supplementation of geranylgeraniol in three patients with MVK deficiency/HIDS. The study was approved by the Ethical Committee, Motol University Hospital, Prague, Ref. Number EK 30/22, and by the Czech Agriculture and Food Inspection Authority who oversees the field of food supplements. All patients signed informed consent with the study.

### Dietary supplementation

Geranylgeraniol in a form of dietary supplement GG Pure (Extendlife Natural Products) containing GG gold®30 Annatto Extract, 500 mg capsule (30% geranylgeraniol, 150 mg per capsule) was administered once daily for 3 months.

### Patients

A total of three patients were involved in the study, two females and one male, age of 20-30, and 50-60. All patients were compound heterozygotes for mutations in MVK. We included patients who are followed in Motol hospital and are coming for regular visits. Inclusion criteria required the subjects to be above 12 years old, confirmed diagnosis of HIDS and compliance with the study. Patients were treated with NSAIDs and Anakinra on demand at the time of the attack. Patient 1 used Anakinra only for more severe attacks. She started the study and used GG supplementation for 6 weeks when she experienced an allergic reaction to a combination of non-steroid anti-inflammatory drugs taken together with GG supplement. Subsequent investigation failed to prove the causality of the reaction, but the patient stopped the supplement during the rest of the pilot period. Patient 2 applied Anakinra regularly every 1-2 days and was on maintenance corticosteroid therapy (Prednisone 10 mg every other day) in addition to GG supplementation. Patient 3 used GG supplementation throughout the study and did not apply Anakinra during the study period. At the time of sampling, all patients were in a quiet period, away from inflammatory attacks, without acute inflammatory symptoms. Statin group consisted of three controls, two females aged 50-60 and one male 40-50 years, all were using Atorvastatin at a dose of 10 mg/day.

## Plasma and serum collection

Blood was collected into ethylenediaminetetraacetic acid (EDTA) tubes at room temperature and centrifuged at 3000 rpm for 5 minutes. Plasma was immediately frozen at -80°C and stored until profiling. Serum was similarly obtained from a routine collection of coagulable blood.

## Liver function assessment/enzyme assays

Aspartate aminotransferase (AST) and alanine aminotransferase (ALT) were assessed by the routine colorimetric IFCC methods in serum. Bilirubin levels in serum were measured by vanadate oxidase method.

## Lipid measurements

Lipid profile in serum consisting of total cholesterol, HDL-cholesterol (HDL), LDL-cholesterol (LDL) and triglycerides (TAG). Lipids were measured by routine methods using enzyme CHOD-PAP end point test (Enzymatic Colorimetric Determination in sera).

## Flow cytometry measurements

Flow cytometry for B panel analysis was performed from isolated PMBCs, using the following panel of antibodies - dried mixture of IgD FITC, CD27 PE, CD24 PerCP-Cy5.5, CD19 PE-Cy7, CD21 APC, and CD38 APC-Cy7 (Custom-design dry reagent tube, Exbio Praha, Vestec, Czech Republic) that allowed to determine subsets of B cells defined as naïve, MZ-like, switched memory B cells and plasmablasts (see Fig. S2 for gating strategy).

## Analysis of isoprenoid pyrophosphates in plasma

### LC-MS/MS analysis

Negative-ion electrospray ionization (ESI) LC-MS/MS analysis of isoprenoid pyrophosphates (GPP, FPP, and GGPP) in plasma and added ^15^N_5_-ADP internal standard was conducted on an ABI-4000 QTRAP triple stage mass spectrometer, coupled to a Shimadzu 20 ADX HPLC system with a SIL-20AC autosampler. To separate the three analysts and the internal standard, a Waters Atlantis dC18 HPLC column (4.6 x 150 mm, 100 Å, 3 µm) was used for a linear gradient separation starting with 40% mobile phase A (5 mM NaHCO_3_ in water) to 100 % mobile phase B (1% NH_4_OH in 1:1 Methanol/acetonitrile) in 5 min at the flow rate 1.0 ml/min. Multiple reaction monitoring (MRM) was used for detection of GPP (313.1 → 79.0), FPP (381.1 → 79.0), GGPP (449.2 → 79.0), and ^15^N_5_-ADP standard (431.0 → 79.0). An aliquot of 5 µL of each sample as prepared below was injected and analyzed twice. The resultant data from two injections were averaged.

### Sample preparation

To prepare samples for MRM LC-MS/MS analysis, plasma samples (200 µL each in an Eppendorf tube) were added 800 µL of methanol containing 2 µg of ^15^N_5_ adenosine diphosphate (ADP) as an internal standard. The samples were vortexted, and centrifuged to precipitate the protein. The supernatant was transferred to a new vial, dried under a stream of nitrogen at room temperature, and reconstituted in 100 µL methanol. A set of four-point calibration samples with 1 ng, 5 ng, 10 ng, and 20 ng of each analyte in 900 µL with the same amount of the internal standard were also prepared to establish a linear calibration curve for absolute quantification of the three analytes.

## Plasma proteomic profiling by SomaScan 7K assay

For each sample, 500 μL of frozen plasma was shipped on dry ice to the Genome Technology Access Center (GTAC) core facility at Washington University in St. Louis for high-density protein expression analysis via SomaScan assay^18^ (somalogic). Profiles of 7290 analytes were acquired. Pre-processing was performed by GTAC core facility at Washington University in St. Louis: raw relative fluorescence units measurements for every SOMAmer reagent were normalized subsequently with hybridization normalization, plate scaling, median scaling and calibrator normalization and transformed in log_2_ scale. Subsequently, differential analysis was performed using limma statistics implemented in web-based platform Phantasus version: 1.21.5^19^. The probes recognizing the same analytes were not collapsed as they can recognize different activation states of the same proteins, or different domains/subunits of the same proteins (further specified in SomaScan assay annotation **Supplementary Table 1**). Gene set enrichment analysis (GSEA) was performed on proteins ranked by limma statistics. To determine enriched pathways either all differentially detected proteins were used or 200 most upregulated and 200 most downregulated proteins were used in MSigDB CP: canonical pathways and H: hallmark gene sets collections.

## Targeted detection of proteins

Proteins in plasma were determines by ELISA using following kits: Human Proteinase 3 (PRTN3) ELISA, (BioVendor, #RAI001R, Czech Republic); LEGEND MAX™ Human Myeloperoxidase ELISA Kit (Biolegend, #440007 USA) as recommneded by manufacutrer’s protocols. Proteins in serum: serum amyloid A (SAA), C-reactive protein (CRP) and IgD were analyzed by routine laboratory methods, SAA by immunoturbidimetry, CRP and IgD by nephelometry.

## Statistical analysis

If not stated otherwise data represents mean ± SEM from N samples as indicated. *P* values were determined using statistical methods described in figure legends using GraphPad Prism v10.0.2, statistical significance was defined as p < 0.05.

## Results

### Study design and rationale

Key metabolites synthesized through mevalonate pathway include non-sterol isoprenoids, which are crucial for protein prenylation such as farnesylation and geranylgeranylation (**Fig. 1A**). The defective protein prenylation, presumably due to the shortage in isoprenoid intermediates, has been observed in monocytes from patients with MVK deficiency and connected to inflammation in HIDS^20^. We aimed to test efficacy and safety of supplementation of isoprenoid end product (**Fig. 1B**), geranylgeraniol (GG; Gold®30 Annatto Extract 500 mg, 30% geranylgeraniol, 150 mg), administered for 3 months (**Fig. 1C**). The plasma and PBMC from HIDS patients were collected before and after 3 months of the treatment. To determine safety of the treatment, additional samples were collected at 1 month of the treatment for routine investigation of blood counts and hepatic function. We also included a control group of healthy donors and group of donors using statins to compare HIDS condition with pharmacologically inhibited mevalonate pathway (**Fig. 1D**). Three patients with HIDS were enrolled in pilot study (two females and one male, aged 20-30 and 50-60, all compound heterozygotes for mutations in MVK). Patients were treated with NSAIDs and Anakinra on demand at the time of attack; the patient 2 applied Anakinra regularly every 1-2 days and was on maintenance corticosteroid therapy, in addition to GG supplementation (**Fig. 1E, F**). The subjective clinical status was followed by questionnaire with 6 relevant parameters – overall feeling, temperature, fatigue, skin presentation, headache and abdominal pain, the objective clinical presentation was checked during entry and follow-up visits by physicians. Despite detailed instructions on monitoring their clinical status, the patients did not fill in the questionnaires consistently. The results are therefore inconclusive, but showed stabilization to slight improvement, most notably in general fatigue. All patients took the dietary supplement at the recommended dose of one capsule per day (150 mg of geranylgeraniol), without significant side effects with exception of one patient with a suspected allergic reaction at the second month of use, which was more likely to the concomitant non-steroidal anti-inflammatory drugs and mild, short, and self-limiting muscle pain reported by another patient.

**Figure 1.**
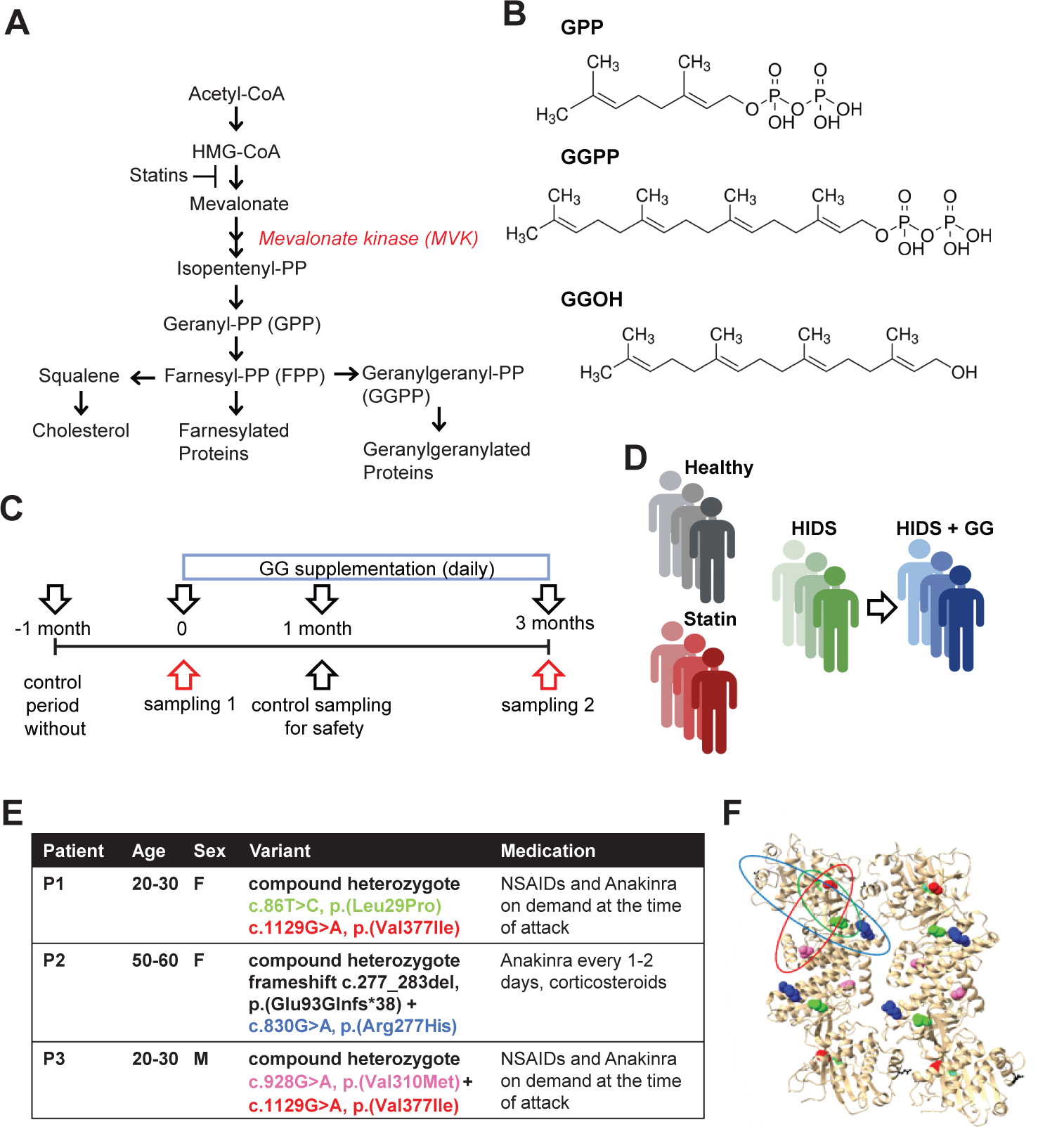
Pilot study design. **A)** Scheme of the mevalonate pathway. **B**) Structures of metabolites and geranylgeraniol (GGOH) as indicated. **C**) Scheme of geranylgeraniol (GG) supplement administration. GG was administered as nutritional supplement for 3 months to patients with HIDS. **D**) Scheme of groups included in the study. **E**) Involved HIDS patient information. NSAIDs, non-steroidal anti-inflammatory drugs. **F**) MVK ribon structure indicating mutations in HIDS patients involved in the study; color coding as in E.

### GG supplementation does not induce liver toxicity, nor does it alter lipid composition and isoprenoid pyrophosphate levels in plasma

To evaluate the effect of the GG supplementation on liver functions we assessed activity of aspartate transaminase (AST), alanine transaminase (ALT) and serum levels of bilirubin. The normal values of these parameters are 0.16-0.72 μkat/L for AST, 0.17-0.78 μkat/L for ALT and 5.0-21.0 μmol/L for bilirubin. At any timepoint of the GG supplementation the values did not exceed the limits, except minor increase in patient 2 in AST at 1 month of supplementation that normalized at month 3 (**Fig. 2A**). To evaluate the impact of GG supplementation on lipid metabolism, we analyzed cholesterol, high-density lipoprotein (HDL), and low-density lipoprotein (LDL) levels in patient plasma at baseline (time 0), as well as after 1 and 3 months of GG supplementation. No significant differences in any of these assessed parameters were found following GG supplementation (**Fig. 2B**). This data indicates that GG used in a given dose does not interfere with cholesterol and lipid transport pathways in HIDS patients. Next, the rationale behind GG supplementation lies in its potential to replenish geranylgeranyl moieties possibly depleted due to the MVK deficiency. To investigate whether GG can contribute to the plasma pool of isoprenoid pyrophosphates, we examined the plasma levels of geranyl pyrophosphate (GPP), farnesyl pyrophosphate (FPP), and geranylgeranyl pyrophosphate (GGPP) using targeted LC-MS/MS analysis. Among the tested isoprenoid intermediates, only GGPP exhibited a significant decrease in HIDS patients compared to controls (**Fig. 2C, Fig. S1**). As anticipated, GGPP levels were notably reduced also in the statin group, indicating that GGPP is the intermediate most susceptible to depletion in the settings of MVK pathway inhibition. However, supplementation with GG did not result in increase in GGPP levels suggesting that geranylgeraniol may not be readily metabolized to pyrophosphate in the circulation or undergoes metabolic turnover that precludes its detection in the plasma at this dose.

**Figure 2.**
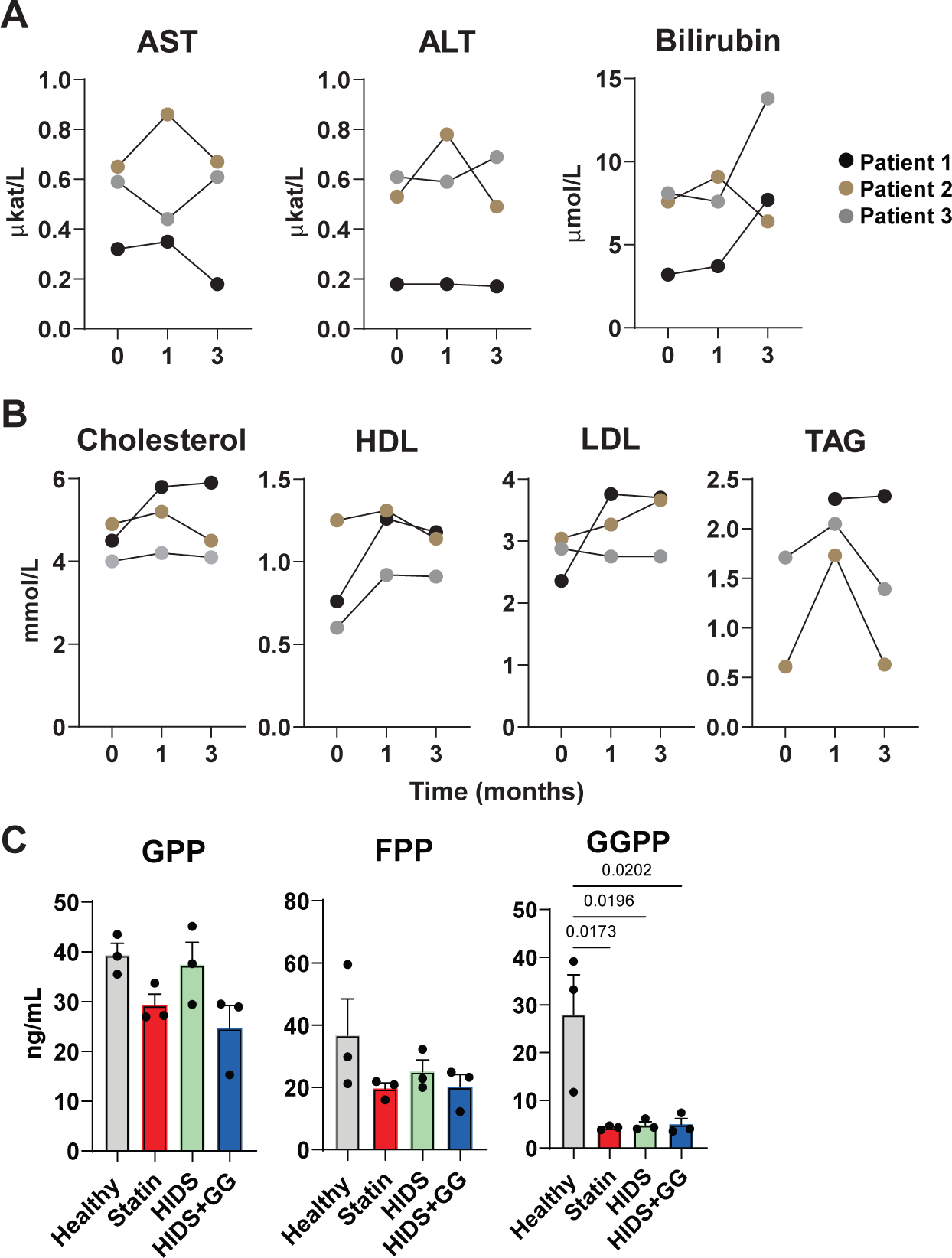
GG supplementation does not exert liver toxicity and does not alter lipid and isoprenoid plasma composition. **A**) Assesment of the liver function by measurement of enzyme activity (AST and ALT) and bilirubin levels in serum at indicated times of GG supplementation. **B**) Lipid measurements in plasma in three patients at timepoints post GG supplementation as indicated. **C**) Isoprenoid pyrophosphate levels in plasma analyzed by LC-MS/MS. GPP, geranyl pyrophosphate; FPP, farnesyl pyrophosphate; GGPP, geranylgeranyl pyrophosphate. Data in (C) represent mean ± SEM. *P* values were determined using repeated measure one-way ANOVA with Sikad’s test (A, B); and one-way ANOVA with Tuckey’s test (C).

### Phenotyping of B lymphocytes

The B lymphocytes were determined as CD19^+^ cells in peripheral blood mononuclear cells (PBMCs) from all experimental groups. The absolute B cell counts were normal in all samples, ranging from 0,04 to 0,15 (normal values 0,03-0,40 x 10^9^/L) and the total counts did not change after GG supplementation (data not shown). In addition, subpopulations of B lymphocytes defined as naΪve, MZ like lymphocytes, memory, switched memory and plasmablasts were detected using the B cell panel (as described in *Methods* section). The samples post-treatment with GG trended towards increased CD19^+^ B cells and decreased plasmablasts in patients whose initial plasmablast counts were higher than in controls (**Fig. 3, Fig, S2**). However, the results are strongly influenced by the small number of patients and also by the lack of initial B panel determination in patient 2. This trend needs to be verified in a targeted follow-up in a larger cohort of patients with MVK deficiency.

**Figure 3.**
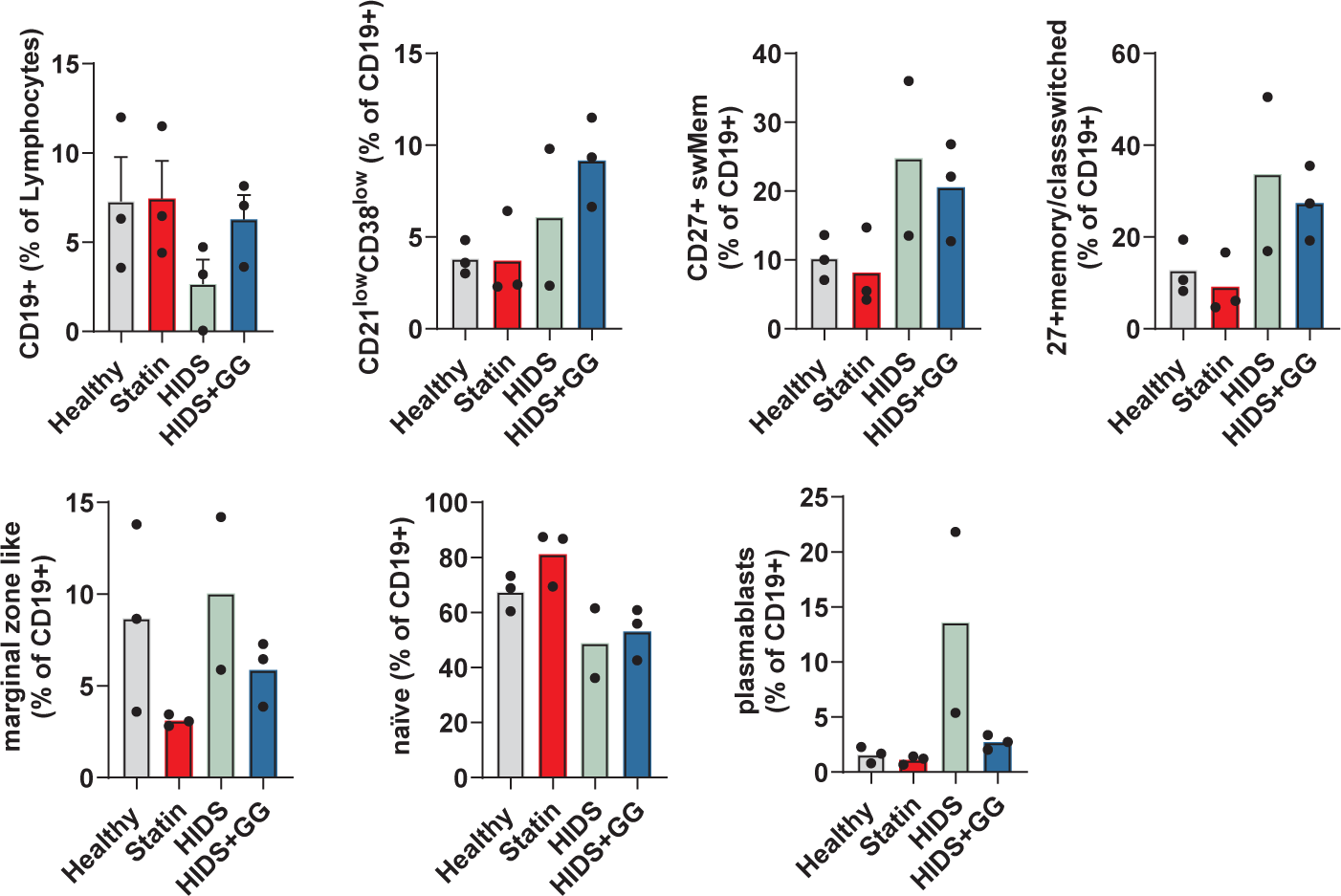
B lymphocyte analysis. Cells from PBMCs were analyzed by flow cytomery. Where N = 3, data are mean ± SEM, *P* values were determined using one-way ANOVA with Tuckey’s test.

### GG supplement reverses HIDS-specific plasma protein signature

To examine global proteomic alterations in patient plasma, we utilized SomaScan 7K assay, identifying approximately 7290 analytes. Plasma from all experimental groups (**Fig. 1D**) was analyzed in one run. Principal component analysis (PCA) based on all detected analytes showed separation between healthy control donors and samples from HIDS patients, indicating a unique plasma protein profile associated with HIDS (**Fig. 4A**). Statin group was less separated from healthy control group and further distinguished from HIDS. GG supplementation induced alteration in the plasma protein composition in HIDS but did not lead to normalization of the global protein profile. We aimed to utilize this unique rare disease dataset to get an insight into specific plasma changes associated with HIDS. Although the plasma from HIDS patient show altered protein composition, individual gene differences did not reach statistical significance due to limited sample size and inclusion of patient with clearly different inflammatory status (patient 2) resulting from her intensive and continuous anti-inflammatory therapy consisting of IL-1 blockade and corticosteroids (**Fig. 4B, Fig. S3A, B**). Since pathway analysis provides a broader biological context by uncovering coordinated changes in groups of genes/proteins, we performed pathway analysis focusing on the upregulated and downregulated proteins detected in HIDS plasma compared to healthy controls. The analysis revealed an associations with critical processes including post-translational modification, innate immune response, cytokine signaling, and geranylgeranylation (**Fig. 4C Fig. S3C**). Among downregulated pathways we found pathways connected to signaling and extracellular matrix organization. Statin group at this size did not show extensive difference when compared with healthy control group, however, pathway analysis suggested enrichment in Rho signaling indicating potential intersection of the processes involved in MVK block by statins and dysfunction in HIDS patients involving small GTPases (**Fig. S3D, E**). These findings align with anticipated infammatory status of the HIDS patients and abnormalities likely stemming from the deficiency of isoprenoids.

**Figure 4.**
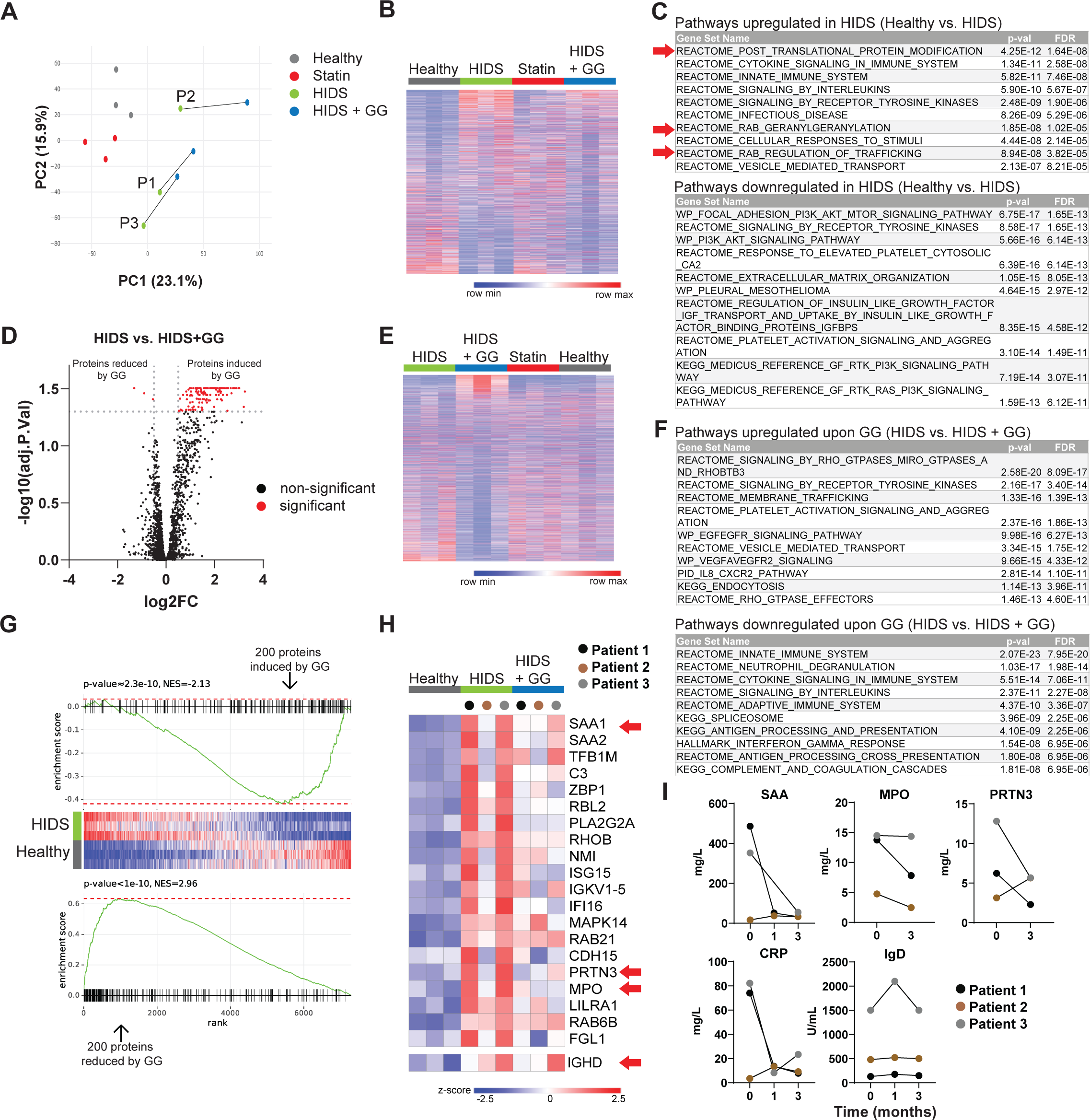
GG supplement reverses HIDS-associated protein plasma signature. **A**) PCA plot of SomaScan plasma proteomics including 7289 targets. Patients 1-3 are indicated in the plot. **B**) Comparison of proteins in ‘Healthy’ and ‘HIDS’ groups using limma, proteins were sorted based on t statistics. **C**) Pathway analysis of 200 most increased and 200 most decreaed proteins in ‘HIDS’ vs. ‘Healty’ compared as in B. Red arrows highlight pathways indicating involvement of GTPase signaling. **D**) Volcano plot of proteomic data comparing ‘HIDS‘ vs. ‘HIDS + GG‘. Significanly different proteins as determined by adjPval (0.05; correction for multiple testing using the Benjamini–Hochberg method) and log fold change (FC) 0.5 are shown in red. **E**) Comparison of proteins in ‘HIDS’ vs. ‘HIDS + GG’ groups using limma, proteins were sorted based on t statistics. **F**) Pathway analysis of 200 most increased and 200 most decreased proteins in ‘HIDS’ vs. ‘HIDS + GG’ compared as in E. **G**) GSEA statistics comparing HIDS protein signature and effect of GG in HIDS patients (200 most upregulated and 200 most downregulated proteins form ‘HIDS’ vs. ‘HIDS + GG’ were used). **H**) 20 most upregulated proteins from comparison between ‘Healthy’ and ‘HIDS’ including only samples from patients P1 and P3. IgD expression is shown separately. Red arrows indicate targets chosen for *in vitro* validation. **I**) Targeted detection of proteins in plasma or serum of HIDS patients at indicated time of treatment with GG.

Next we focused on differences in HIDS plasma induced by GG. We found 140 proteins significantly altered by GG supplementation (137 up a 3 down; **Fig. 4D**). Pathway enrichment analysis highlighted processes centering around Rho GTPase signaling, tyrosine kinase receptor signaling, and membrane trafficking upon GG treatment (**Fig. 4E, F**). This suggested a potential interference of GG supplement with protein prenylation-associated signaling. The most upregulated proteins were specifically up in GG-treated group and reached up to 3 log2 difference in intensity units compared to healthy donors (**Fig. S4A**). To investigate whether GG has a potential to modulate the characteristic protein profile of HIDS patient plasma, we ranked proteins based on the comparison between controls and HIDS samples and performed Gene Set Enrichment Analysis (GSEA) using proteins regulated by GG. The GSEA analysis indicated a statistically significant overlap between proteins elevated in HIDS and proteins downregulated by GG, as well as proteins decreased in HIDS and upregulated by GG (**Fig. 4G, Fig. S4B**). Similarly, when proteins were ranked based on comparison between HIDS and HIDS + GG groups, GSEA showed significant overlap with HIDS disease signature (**Fig. S4C**). These results suggests that GG supplementation has potential to revert key features of HIDS protein signature in patient plasma.

To experimentally validate changes in inflammatory signature of HIDS patients upon GG treatment we aimed to use the proteomics dataset to find suitable markers. For this purpose, we compared healthy controls with HIDS patients including only patient 1 and 3, excluding patient 2 who was treated by consistent corticosteroid treatment (**Fig. 4H**). We selected several protein targets that showed downregulation/normalization by GG and validated their expression by targeted standard laboratory tests/ELISA assays. The following proteins were evaluated: Serum amyloid A (SAA), Myeloperoxidase (MPO) and Proteinase 3 (also Myeloblastin; PRTN3). We also included IgD, which is one of the diagnostic markers of HIDS and C-reactive protein (CRP) which is a part of routine laboratory evaluation (although IgD did not show change upon GG and CRP was not among the most increased proteins in HIDS) (**Fig. 4H**). Consistent with proteomic data SAA and MPO showed decrease in patients upon GG supplementation for 1 or 3 months (**Fig. 4I, Fig S4D, E**). PRTN3 dropped in two patients and increased in patient 2. Consistent with plasma proteomics, IgD levels remained unchanged. CRP followed the trend at SAA and showed decrease in patients 1 and 3. Therefore, SAA, MPO, CRP could be potentially utilized as markers of the GG efficacy in HIDS patients, however more data must be collected to validate a robustness of the effect on these markers. Collectively, these findings indicate that GG supplementation has a potential to change the altered plasma protein signature associated with HIDS and provide a starting point for future larger and more targeted studies.

## Discussion

Clinical symptoms in patients with MKD/HIDS are dominated by immune activation *(for recent update, UpToDate2024,* https://www.uptodate.com/contents/hyperimmunoglobulin-d-syndrome-clinical-manifestations-and-diagnosis), manifested as febrile episodes accompanied by other inflammatory symptoms, all of which are typical of autoinflammatory diseases associated with inflammasome activation.

Current therapeutic approaches in MKD/HIDS focus on symptomatic management by suppressing inflammation. This includes medications like colchicine, non-steroidal anti-inflammatory drugs (NSAIDs), and cytokine blockade targeting TNF-α and IL-1^20,21,^. However, these options have limited efficacy and long-term sustainability, and do not address the underlying metabolic defect.

In this small pilot study, we introduce a novel approach targeting the metabolic basis of MKD/HIDS. This approach investigates the supplementation of metabolites downstream of the enzymatic block with isoprenoid substitute, specifically geranylgeraniol. The strategy aims to bypass the metabolic deficiency and potentially mitigate downstream inflammatory sequelae.

No significant changes were observed in the lipid profiles of the patients following GG administration. This aligns with previous reports by Simon et al.^22^, which demonstrated normal cholesterol levels in MKD/HIDS individuals. Therefore, the lack of alteration in baseline cholesterol and triglyceride levels was unsurprising.

As anticipated, the comprehensive analysis of plasma proteins in HIDS patients suggested enhancements in various pathways linked to inflammation and immune signaling. The variability in the protein profiles of HIDS patients correlated with their inflammatory status, as demonstrated by the diminished inflammation in one patient treated with corticosteroids, supported by the expression of acute inflammatory markers such as SAA and CRP (**Fig. 4H**, Interestingly, the global protein analysis also detected enrichment in pathways involved in post-translational protein modifications, Rab signaling, and membrane trafficking. Accumulation of unprenylated Rab proteins has been previously detected in cells with decreased MVK activity^23^. However the authors showed that while decrease in Rho/Rac/Rap-family GTPases lead to production of IL-1β in human cells, targeting Rab prenylation had no effect on production of this cytokine, suggesting potentially distinct function of Rab geranylgeranylation. Conversely, the pathways downregulated in HIDS were primarily associated with the regulation of the extracellular matrix. In this regard, previous studies have linked geranylgeraniol supplementation with changes in serum collagen levels in mice^24^.

The proteomic analysis revealed significant alterations in the protein profiles of patients solely upon GG supplementation, without any additional interventions or adjustments to their existing therapy. Significant changes in protein expression (140 proteins) upon the GG can serve as an indicator of the patients’ adherence to the treatment. The pathways identified predominantly centered on signaling through small GTPases and membrane trafficking, indicating that GG holds promise as a strategy to modulate these processes in humans.

Key finding of our study is that GG supplementation demonstrated the ability to reverse the protein signature in HIDS patients as indicated by proteomics and confirmed by targted measurements. Among the candidate proteins responsive to GG treatment are numerous key immunoregulatory proteins such as SAA, Ubiquitin-like protein ISG15 (ISG15), Leukocyte immunoglobulin-like receptor subfamily A (LILRA), C-X-C motif chemokine 13 (CXCL13), Ig Kappa chain V-I region HK102-like (IGKV1-5), Myeloperoxidase (MPO), Myeloblastin (PRTN3) and others. We validated expression of MPO and PRTN3 which are connected to neutrophil activation that might have implication in inflammasomopathies^25,26^. These targtes can serve as potential markers for assessing the efficacy of GG supplementation in more targeted studies in future. The IgD levels remained unaffected by the GG supplement supporting the notion that elevated IgD level might not be directly associated with the inflammatory status in HIDS^12,27^. Although GG treatment induced changes in many HIDS-associated proteins, it did not fully normalize the global plasma profile (**Fig. 4A**). This is likely driven by the strong GG-dependent protein signature that is irrespective of the HIDS status (such as in **Fig. S4A**). In future studies, including healthy group supplemented by GG would help to further determine extent of the effect of GG outside of the inflammatory context of HIDS. Of note, we did not detect changes in classical inflammatory cytokines such as IL-1β, IL-6 or TNF in plasma proteomics.

Despite the observed effectiveness of the GG supplement in altering prenylation-associated pathways, the levels of isoprenoid intermediates (GPP, FPP, GGPP) remained unchanged following GG supplementation at the dosage used (see **Fig. 2**). Geranylgeraniol can be converted endogenously in polyprenol salvage pathway and utilized for protein prenylation^28,29^. As such, geranylgeraniol is known to reverse defects in protein prenylation in various biological contexts and mitigate secondary effects of several drugs. For example, it is known to reduce statin-induced cytotoxicity through the activity of geranylgeranyl transferases (GGTs)^30,31^. However, whether geranylgeraniol increases the levels of GGPP has not been conclusively demonstrated to our knowledge. Intriguingly, the enzymatic machinery facilitating this process remains incompletely understood, as only a few enzymes (kinase and phosphate kinase) necessary for conversion of geranylgeraniol to GGPP have been identified in plants and cyanobacteria^28^. Among other mechanisms, geranylgeraniol can also influence the expression of small GTPases, which may be linked to the plasma signature observed in GG-treated HIDS patients in this study^32^. Interestingly, it has been shown that endogenous geranylgeraniol production in macrophages contributes to the establishment of endotoxin tolerance^33^. Our data suggests that at the given dose, geranylgeraniol does not replenish the levels of GGPP in the circulation, undergoes turnover that precludes the capture of changes in plasma, or acts directly in the form of alcohol. As such, in our study GGPP did not serve as a suitable indicator of the GG supplementation in plasma. However, we assume that the strong significant change in protein expression upon GG detected by proteomics can serve as an indicator of GG activity and as a basis for assessing the GG activity in future studies.

A primary limitation of this pilot study investigating GG supplementation in MKD/HIDS patients is the restricted sample size due to the rarity of the disease. Expanding the patient cohort in future studies will require collaborations with other international centers, leveraging international networks and patient registries. Another limitation is the dosage used, which is based on the manufacturer’s recommendation in the indication for supplementation with statins therapy. This regimen was selected as safe for the pilot study but is likely inadequate in the absence of isoprenoids in MVK deficiency. In this context, the minimum effective dose administered in a given situation in the mouse model was several times greater^17^. Further studies in larger patient cohorts will be required to optimize the dosing scheme. To facilitate such future expansion, the current pilot study aimed to identify simpler and more readily available biomarkers to monitor the therapeutic efficacy of GG substitution beyond complex proteomic profiling. Neutrophil antigens readily assessed using standard ELISA assays were chosen as potential candidates for this purpose.

## Conclusion

The primary safety objective of the study was successfully met. Aside from the described allergic reaction, which was deemed unrelated to the GG preparation itself, no adverse effects were observed following GG supplementation. The clinical response was modest, with some patients experiencing a slight reduction in the frequency of inflammatory attacks and slightly improved overall felling and fatigue. While the administered dose and three-month study duration might have been suboptimal, this observed, even if mild, improvement is encouraging. Changes in laboratory parameters provided further evidence for the potential efficacy of GG supplementation in MKD/HIDS. The observed shifts in the proteomic profile towards an anti-inflammatory state suggest that targeting this pathway could offer long-term benefits for disease management. Validation of the dynamics of neutrophil targets as a markers of substitution efficacy holds promise for facilitating larger and international clinical trials to further evaluate this therapeutic strategy in the future.

## Supporting information

Suppelementary Table 1

## Data Availability

All data produced in the present study are available upon reasonable request to the authors.

## Acknowledgements

We thank the Genome Technology Access Center at the McDonnell Genome Institute at Washington University School of Medicine for help with genomic analysis. The Center is partially supported by NCI Cancer Center Support Grant #P30 CA91842 to the Siteman Cancer Center from the National Center for Research Resources (NCRR), a component of the National Institutes of Health (NIH), and NIH Roadmap for Medical Research. This publication is solely the responsibility of the authors and does not necessarily represent the official view of NCRR or NIH. We thank Andrew J. Martin for help with manuscript corrections.

---

**Figure S1.**
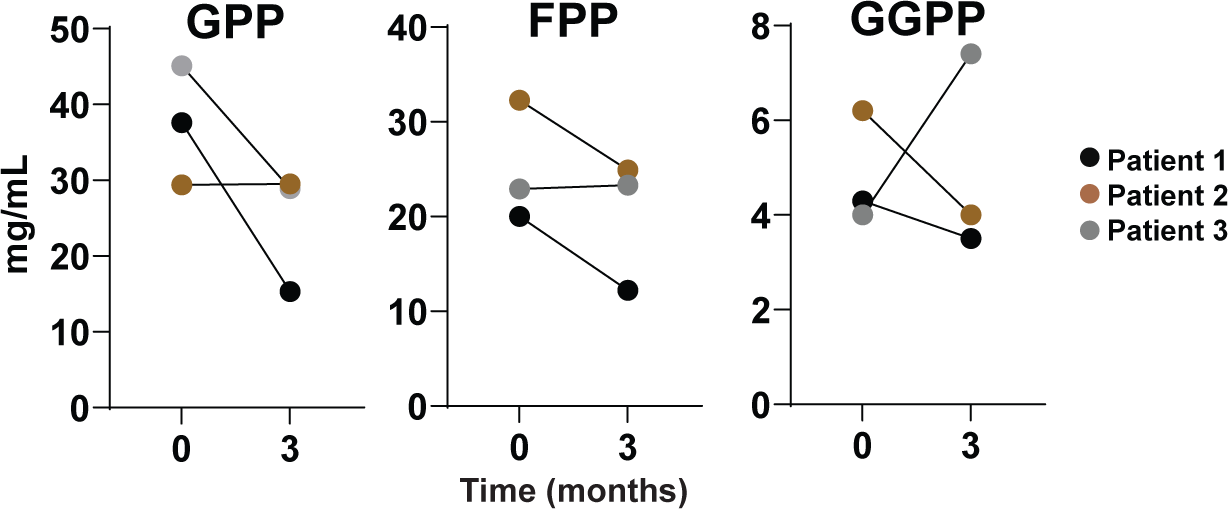
Isoprenoid pyrophosphate levels in plasma analyzed by LC-MS/MS. GPP, geranyl pyrophosphate; FPP, farnesyl pyrophosphate; GGPP, geranylgeranyl pyrophosphate. Data are presented as paired values from patients before and after 3 months of GG treatment. *P* values were determined Wilcoxon matched-pairs signed rant test.

**Figure S2.**
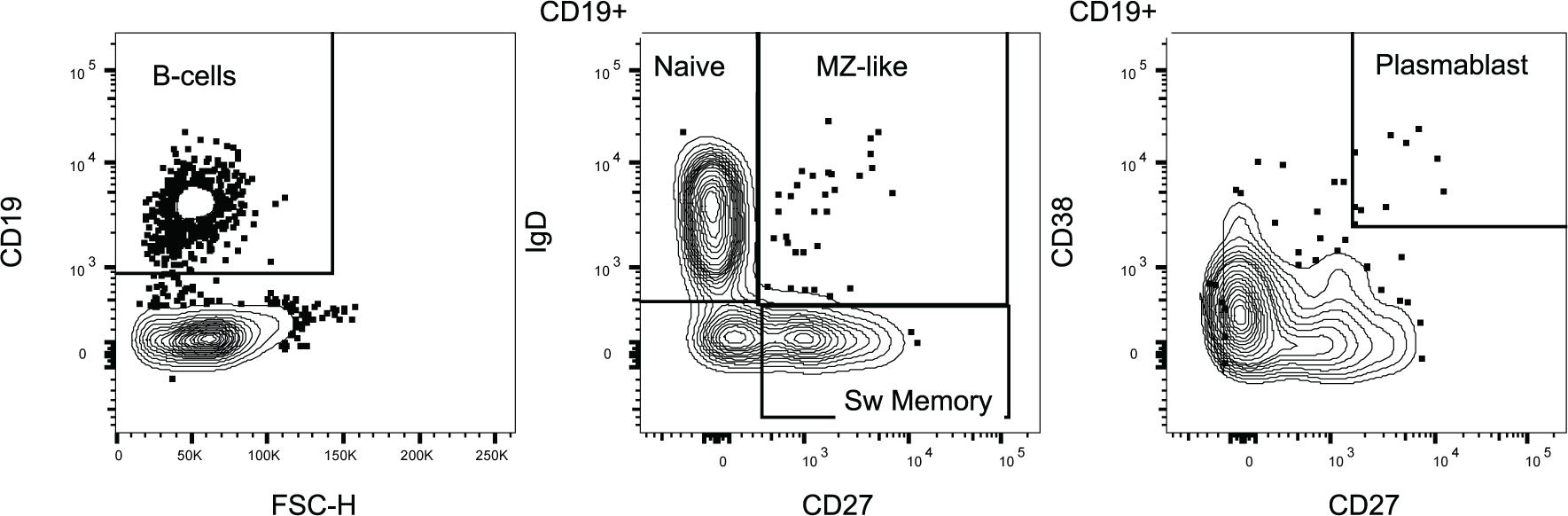
Gating strategy used to identify and resolve B cell populations from PBMCs.

**Figure S3.**
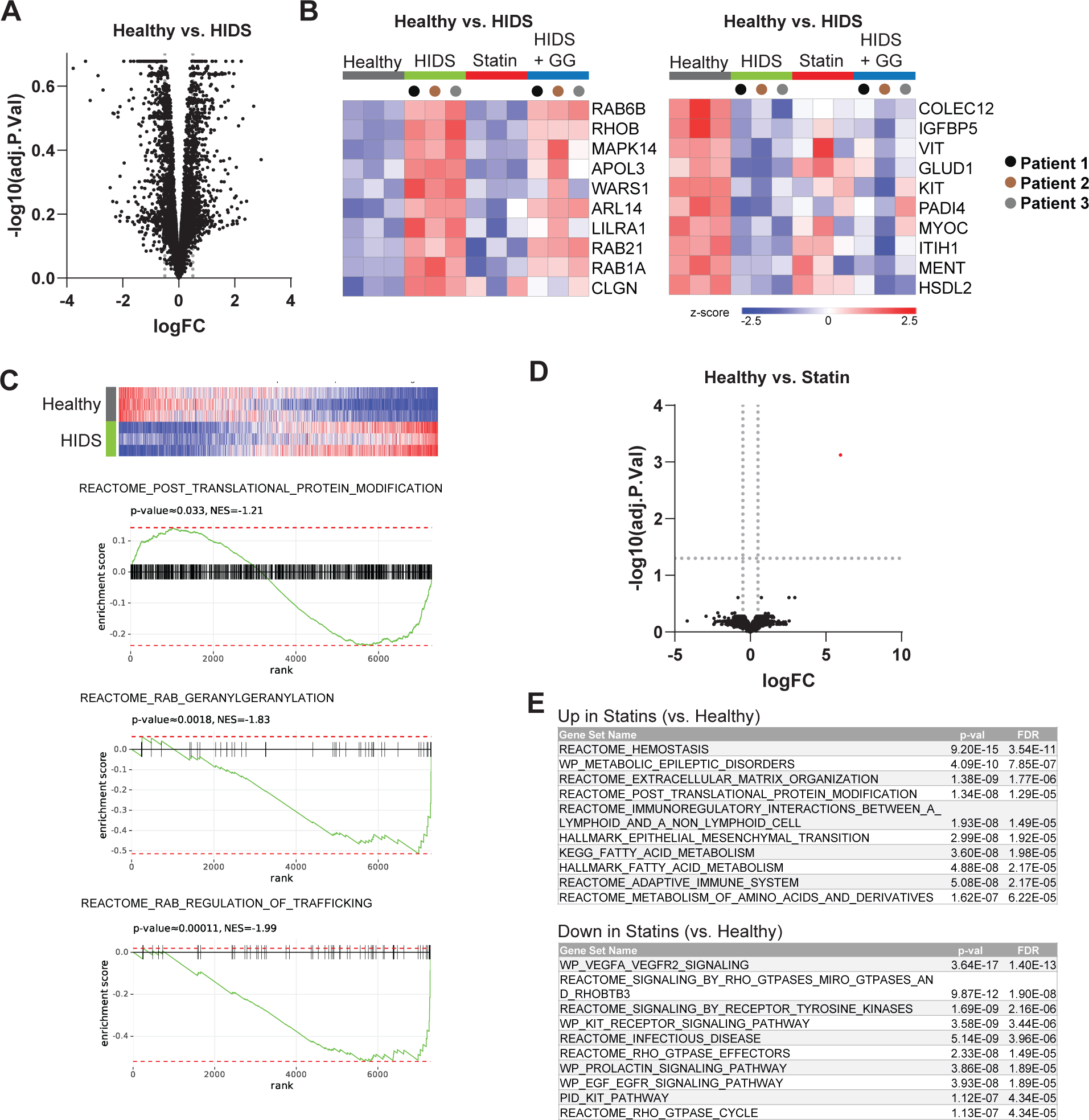
**A**) Volcano plot of proteomic data comparing ‘Helathy’ vs. ‘HIDS’ groups. **B**) Comparison of proteins in ‘Healthy’ and ‘HIDS’ ranked by limma statistics showing 10 most ‘up’ and 10 most ‘down’ proteins detected in HIDS. Individual patiens before and after GG treatment are labeled by color coding. **C**) GSEA statistics comparing ranked by limma ‘Healthy’ vs. ‘HIDS’ protein signature and selected pathways from Fig. 4C. **D**) Volcano plot of proteomic data comparing ‘Healthy’ vs. ‘Statin’ groups. Significanly differential proteins as determined by adjPval (0.05) and log fold change (FC) 0.5 are showed in red. **E**) Pathway analysis of 200 most increased and 200 most decreased proteins in ‘Healthy’ vs. ‘Statin’ compared by limma statistics.

**Figure S4.**
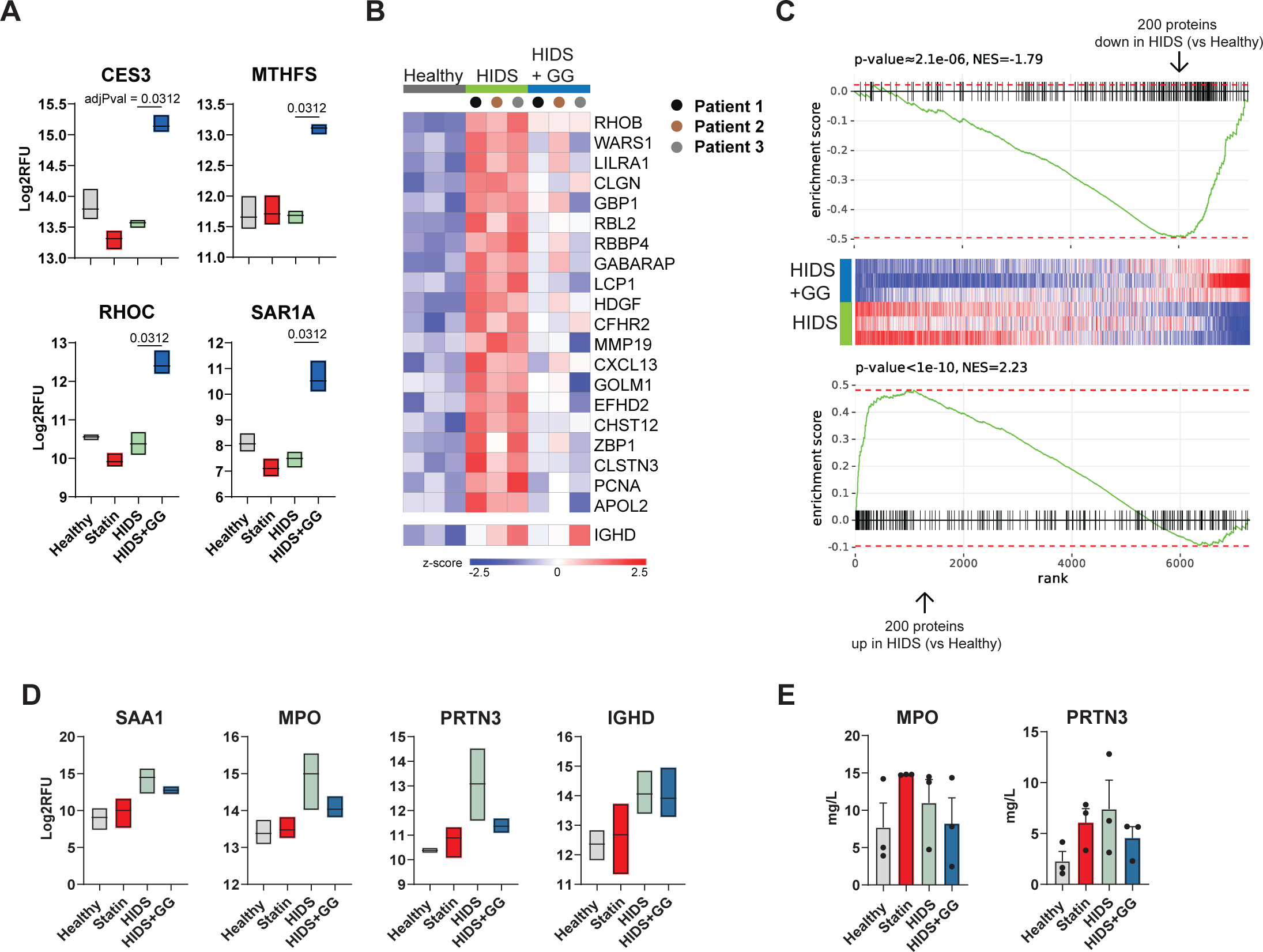
**A)** Example of SomaScan expression data of proteins significantly upregulated by GG in HIDS patients across all experimental groups. **B)** Proteins most increased in ‘Healthy’ vs. ‘HIDS’ ranked by limma statistics. Samples from individual patients are labeled by color coding. **C)** GSEA statistics comparing GG protein signature and 200 most increased and 200 most decreased proteins in ‘HIDS’ compared to ‘Healthy’. **D)** SomaScan data on expression of proteins as indicated. **E)** Protein detection in plasma measured by ELISA. Boxplot in (A) and (D) represent mean/min and max values from N = 3. Adjusted P values in SomaScan data (A) and (D) were determined using the limma package comparing ‘HIDS’ and ‘HIDS + GG‘, and correction for multiple testing using the Benjamini–Hochberg method.

## Notes

### Competing Interest Statement

The authors have declared no competing interest.

### Clinical Trial

NCT06497829

### Funding Statement

This study was funded by internal grant, Motol University Hospital, Prague Czech Republic.

### Author Declarations

The pilot study was approved by the Ethical Committee, Motol University Hospital, Prague, Ref. Number EK 30/22, and by the Czech Agriculture and Food Inspection Authority who oversees the field of food supplements.

## References

(1) Drenth, J. P.; Cuisset, L.; Grateau, G.; Vasseur, C.; van de Velde-Visser, S. D.; de Jong, J. G.; Beckmann, J. S.; van der Meer, J. W.; Delpech, M. Mutations in the Gene Encoding Mevalonate Kinase Cause Hyper-IgD and Periodic Fever Syndrome. International Hyper-IgD Study Group. Nat. Genet. 1999, 22 (2), 178–181. 10.1038/9696.

(2) van der Burgh, R.; Ter Haar, N. M.; Boes, M. L.; Frenkel, J. Mevalonate Kinase Deficiency, a Metabolic Autoinflammatory Disease. Clin. Immunol. 2013, 147 (3), 197–206. 10.1016/j.clim.2012.09.011.

(3) Wang, M.; Casey, P. J. Protein Prenylation: Unique Fats Make Their Mark on Biology. Nat. Rev. Mol. Cell Biol. 2016, 17 (2), 110–122. 10.1038/nrm.2015.11.

(4) Munoz, M. A.; Jurczyluk, J.; Simon, A.; Hissaria, P.; Arts, R. J. W.; Coman, D.; Boros, C.; Mehr, S.; Rogers, M. J. Defective Protein Prenylation in a Spectrum of Patients With Mevalonate Kinase Deficiency. Front. Immunol. 2019, 10.

(5) Masumoto, J.; Zhou, W.; Morikawa, S.; Hosokawa, S.; Taguchi, H.; Yamamoto, T.; Kurata, M.; Kaneko, N. Molecular Biology of Autoinflammatory Diseases. Inflamm. Regen. 2021, 41 (1), 33. 10.1186/s41232-021-00181-8.

(6) Park, Y. H.; Wood, G.; Kastner, D. L.; Chae, J. J. Pyrin Inflammasome Activation and RhoA Signaling in the Autoinflammatory Diseases FMF and HIDS. Nat. Immunol. 2016, 17 (8), 914–921. 10.1038/ni.3457.

(7) Akula, M. K.; Shi, M.; Jiang, Z.; Foster, C. E.; Miao, D.; Li, A. S.; Zhang, X.; Gavin, R. M.; Forde, S. D.; Germain, G.; Carpenter, S.; Rosadini, C. V; Gritsman, K.; Chae, J. J.; Hampton, R.; Silverman, N.; Gravallese, E. M.; Kagan, J. C.; Fitzgerald, K. A.; Kastner, D. L.; Golenbock, D. T.; Bergo, M. O.; Wang, D. Control of the Innate Immune Response by the Mevalonate Pathway. Nat. Immunol. 2016, 17 (8), 922–929. 10.1038/ni.3487.

(8) Skinner, O. P.; Jurczyluk, J.; Baker, P. J.; Masters, S. L.; Rios Wilks, A. G.; Clearwater, M. S.; Robertson, A. A. B.; Schroder, K.; Mehr, S.; Munoz, M. A.; Rogers, M. J. Lack of Protein Prenylation Promotes NLRP3 Inflammasome Assembly in Human Monocytes. J. Allergy Clin. Immunol. 2019, 143 (6), 2315–2317.e3. 10.1016/j.jaci.2019.02.013.

(9) Bekkering, S.; Arts, R. J. W.; Novakovic, B.; Kourtzelis, I.; van der Heijden, C. D. C. C.; Li, Y.; Popa, C. D.; Ter Horst, R.; van Tuijl, J.; Netea-Maier, R. T.; van de Veerdonk, F. L.; Chavakis, T.; Joosten, L. A. B.; van der Meer, J. W. M.; Stunnenberg, H.; Riksen, N. P.; Netea, M. G. Metabolic Induction of Trained Immunity through the Mevalonate Pathway. Cell 2018, 172 (1–2), 135–146.e9. 10.1016/j.cell.2017.11.025.

(10) Bibby, J. A.; Purvis, H. A.; Hayday, T.; Chandra, A.; Okkenhaug, K.; Rosenzweig, S.; Aksentijevich, I.; Wood, M.; Lachmann, H. J.; Kemper, C.; Cope, A. P.; Perucha, E. Cholesterol Metabolism Drives Regulatory B Cell IL-10 through Provision of Geranylgeranyl Pyrophosphate. Nat. Commun. 2020, 11 (1), 3412. 10.1038/s41467-020-17179-4.

(11) Du, X.; Zeng, H.; Liu, S.; Guy, C.; Dhungana, Y.; Neale, G.; Bergo, M. O.; Chi, H. Mevalonate Metabolism–Dependent Protein Geranylgeranylation Regulates Thymocyte Egress. J. Exp. Med. 2019, 217 (2), e20190969. 10.1084/jem.20190969.

(12) Ammouri, W.; Cuisset, L.; Rouaghe, S.; Rolland, M.-O.; Delpech, M.; Grateau, G.; Ravet, N. Diagnostic Value of Serum Immunoglobulinaemia D Level in Patients with a Clinical Suspicion of Hyper IgD Syndrome. Rheumatology (Oxford). 2007, 46 (10), 1597–1600. 10.1093/rheumatology/kem200.

(13) Maity, P. C.; Blount, A.; Jumaa, H.; Ronneberger, O.; Lillemeier, B. F.; Reth, M. B Cell Antigen Receptors of the IgM and IgD Classes Are Clustered in Different Protein Islands That Are Altered during B Cell Activation. Sci. Signal. 2015, 8 (394), ra93. 10.1126/scisignal.2005887.

(14) Silva, R. C. M. C. Hyper-IgD Syndrome: Caused by Deficiency on Ras Prenylation and Trained Immunity? J. Clin. Immunol. 2023. 10.1007/s10875-023-01548-x.

(15) Morgan, M. A.; Sebil, T.; Aydilek, E.; Peest, D.; Ganser, A.; Reuter, C. W. M. Combining Prenylation Inhibitors Causes Synergistic Cytotoxicity, Apoptosis and Disruption of RAS-to-MAP Kinase Signalling in Multiple Myeloma Cells. Br. J. Haematol. 2005, 130 (6), 912–925. 10.1111/j.1365-2141.2005.05696.x.

(16) Munoz, M. A.; Skinner, O. P.; Masle-Farquhar, E.; Jurczyluk, J.; Xiao, Y.; Fletcher, E. K.; Kristianto, E.; Hodson, M. P.; O’Donoghue, S. I.; Kaur, S.; Brink, R.; Zahra, D. G.; Deenick, E. K.; Perry, K. A.; Robertson, A. A.; Mehr, S.; Hissaria, P.; Mulders-Manders, C. M.; Simon, A.; Rogers, M. J. Increased Core Body Temperature Exacerbates Defective Protein Prenylation in Mouse Models of Mevalonate Kinase Deficiency. J. Clin. Invest. 2022, 132 (19). 10.1172/JCI160929.

(17) Marcuzzi, A.; Pontillo, A.; De Leo, L.; Tommasini, A.; Decorti, G.; Not, T.; Ventura, A. Natural Isoprenoids Are Able to Reduce Inflammation in a Mouse Model of Mevalonate Kinase Deficiency. Pediatr. Res. 2008, 64 (2), 177–182. 10.1203/PDR.0b013e3181761870.

(18) Gold, L.; Ayers, D.; Bertino, J.; Bock, C.; Bock, A.; Brody, E. N.; Carter, J.; Dalby, A. B.; Eaton, B. E.; Fitzwater, T.; Flather, D.; Forbes, A.; Foreman, T.; Fowler, C.; Gawande, B.; Goss, M.; Gunn, M.; Gupta, S.; Halladay, D.; Heil, J.; Heilig, J.; Hicke, B.; Husar, G.; Janjic, N.; Jarvis, T.; Jennings, S.; Katilius, E.; Keeney, T. R.; Kim, N.; Koch, T. H.; Kraemer, S.; Kroiss, L.; Le, N.; Levine, D.; Lindsey, W.; Lollo, B.; Mayfield, W.; Mehan, M.; Mehler, R.; Nelson, S. K.; Nelson, M.; Nieuwlandt, D.; Nikrad, M.; Ochsner, U.; Ostroff, R. M.; Otis, M.; Parker, T.; Pietrasiewicz, S.; Resnicow, D. I.; Rohloff, J.; Sanders, G.; Sattin, S.; Schneider, D.; Singer, B.; Stanton, M.; Sterkel, A.; Stewart, A.; Stratford, S.; Vaught, J. D.; Vrkljan, M.; Walker, J. J.; Watrobka, M.; Waugh, S.; Weiss, A.; Wilcox, S. K.; Wolfson, A.; Wolk, S. K.; Zhang, C.; Zichi, D. Aptamer-Based Multiplexed Proteomic Technology for Biomarker Discovery. PLoS One 2010, 5 (12), e15004. 10.1371/journal.pone.0015004.

(19) Kleverov, M.; Zenkova, D.; Kamenev, V.; Sablina, M.; Artyomov, M. N.; Sergushichev, A. A. Phantasus: Web-Application for Visual and Interactive Gene Expression Analysis. bioRxiv 2022, 2022.12.10.519861. 10.1101/2022.12.10.519861.

(20) Politiek, F. A.; Waterham, H. R. Compromised Protein Prenylation as Pathogenic Mechanism in Mevalonate Kinase Deficiency. Front. Immunol. 2021, 12, 724991. 10.3389/fimmu.2021.724991.

(21) Favier, L. A.; Schulert, G. S. Mevalonate Kinase Deficiency: Current Perspectives. Appl. Clin. Genet. 2016, 9, 101–110. 10.2147/TACG.S93933.

(22) Simon, A.; Bijzet, J.; Voorbij, H. A. M.; Mantovani, A.; van der Meer, J. W. M.; Drenth, J. P. H. Effect of Inflammatory Attacks in the Classical Type Hyper-IgD Syndrome on Immunoglobulin D, Cholesterol and Parameters of the Acute Phase Response. J. Intern. Med. 2004, 256 (3), 247–253. 10.1111/j.1365-2796.2004.01359.x.

(23) Jurczyluk, J.; Munoz, M. A.; Skinner, O. P.; Chai, R. C.; Ali, N.; Palendira, U.; Quinn, J. M. W.; Preston, A.; Tangye, S. G.; Brown, A. J.; Argent, E.; Ziegler, J. B.; Mehr, S.; Rogers, M. J. Mevalonate Kinase Deficiency Leads to Decreased Prenylation of Rab GTPases. Immunol. Cell Biol. 2016, 94 (10), 994–999. 10.1038/icb.2016.58.

(24) Chung, E.; Elmassry, M. M.; Cao, J. J.; Kaur, G.; Dufour, J. M.; Hamood, A. N.; Shen, C.-L. Beneficial Effect of Dietary Geranylgeraniol on Glucose Homeostasis and Bone Microstructure in Obese Mice Is Associated with Suppression of Proinflammation and Modification of Gut Microbiome. Nutr. Res. 2021, 93, 27–37. 10.1016/j.nutres.2021.07.001.

(25) Martirosyan, A.; Poghosyan, D.; Ghonyan, S.; Mkrtchyan, N.; Amaryan, G.; Manukyan, G. Transmigration of Neutrophils From Patients With Familial Mediterranean Fever Causes Increased Cell Activation. Front. Immunol. 2021, 12, 672728. 10.3389/fimmu.2021.672728.

(26) Apostolidou, E.; Skendros, P.; Kambas, K.; Mitroulis, I.; Konstantinidis, T.; Chrysanthopoulou, A.; Nakos, K.; Tsironidou, V.; Koffa, M.; Boumpas, D. T.; Ritis, K. Neutrophil Extracellular Traps Regulate IL-1β-Mediated Inflammation in Familial Mediterranean Fever. Ann. Rheum. Dis. 2016, 75 (1), 269–277. 10.1136/annrheumdis-2014-205958.

(27) van der Hilst, J. C. H.; Bodar, E. J.; Barron, K. S.; Frenkel, J.; Drenth, J. P. H.; van der Meer, J. W. M.; Simon, A. Long-Term Follow-up, Clinical Features, and Quality of Life in a Series of 103 Patients with Hyperimmunoglobulinemia D Syndrome. Medicine (Baltimore). 2008, 87 (6), 301–310. 10.1097/MD.0b013e318190cfb7.

(28) Verdaguer, I. B.; Crispim, M.; Hernández, A.; Katzin, A. M. The Biomedical Importance of the Missing Pathway for Farnesol and Geranylgeraniol Salvage. Molecules 2022, 27 (24). 10.3390/molecules27248691.

(29) Bentinger, M.; Grünler, J.; Peterson, E.; Swiezewska, E.; Dallner, G. Phosphorylation of Farnesol in Rat Liver Microsomes: Properties of Farnesol Kinase and Farnesyl Phosphate Kinase. Arch. Biochem. Biophys. 1998, 353 (2), 191–198. 10.1006/abbi.1998.0611.

(30) Jaśkiewicz, A.; Pająk, B.; Litwiniuk, A.; Urbańska, K.; Orzechowski, A. Geranylgeraniol Prevents Statin-Dependent Myotoxicity in C2C12 Muscle Cells through RAP1 GTPase Prenylation and Cytoprotective Autophagy. Oxid. Med. Cell. Longev. 2018, 2018, 6463807. 10.1155/2018/6463807.

(31) Balaz, M.; Becker, A. S.; Balazova, L.; Straub, L.; Müller, J.; Gashi, G.; Maushart, C. I.; Sun, W.; Dong, H.; Moser, C.; Horvath, C.; Efthymiou, V.; Rachamin, Y.; Modica, S.; Zellweger, C.; Bacanovic, S.; Stefanicka, P.; Varga, L.; Ukropcova, B.; Profant, M.; Opitz, L.; Amri, E.-Z.; Akula, M. K.; Bergo, M.; Ukropec, J.; Falk, C.; Zamboni, N.; Betz, M. J.; Burger, I. A.; Wolfrum, C. Inhibition of Mevalonate Pathway Prevents Adipocyte Browning in Mice and Men by Affecting Protein Prenylation. Cell Metab. 2019, 29 (4), 901–916.e8. 10.1016/j.cmet.2018.11.017.

(32) Fliefel, R. M.; Entekhabi, S. A.; Ehrenfeld, M.; Otto, S. Geranylgeraniol (GGOH) as a Mevalonate Pathway Activator in the Rescue of Bone Cells Treated with Zoledronic Acid: An In Vitro Study. Stem Cells Int. 2019, 2019, 4351327. 10.1155/2019/4351327.

(33) Kim, J.; Lee, J. N.; Ye, J.; Hao, R.; Debose-Boyd, R.; Ye, J. Sufficient Production of Geranylgeraniol Is Required to Maintain Endotoxin Tolerance in Macrophages. J. Lipid Res. 2013, 54 (12), 3430–3437. 10.1194/jlr.M042549.

